# Cognitive and emotional effects of bilateral prefrontal anodal tDCS and high-frequency tRNS in schizophrenia: a randomized sham-controlled study

**DOI:** 10.64898/2025.12.25.25342737

**Authors:** Eisa Jafari, Ali Moghadamzadeh, Zahra Vaziri, Akbar Atadokht, Ali Fathi Jouzdani, Roi Cohen Kadosh, Michael A. Nitsche, Daniel M. Blumberger, Mohammad Ali Salehinejad

**Affiliations:** Department of Psychology, Payame Noor University, Tehran, Iran; Department of Educational Sciences and Psychology, University of Tehran, Tehran, Iran; School of Cognitive Sciences, Institute for Research in Fundamental Sciences, Tehran, Iran; Department of Neuroscience and Behavior, Faculty of Medicine of Ribeirão Preto, University of São Paulo, Ribeirão Preto, Brazil; Department of Psychology, University of Mohaghegh Ardabili, Ardabil, Iran; School of Psychology, University of Surrey, Guildford, United Kingdom; Department of Psychology and Neurosciences, Leibniz Research Centre for Working Environment and Human Factors, Dortmund, Germany; Bielefeld University, University Hospital OWL, Protestant Hospital of Bethel Foundation, University Clinic of Psychiatry and Psychotherapy and University Clinic of Child and Adolescent Psychiatry and Psychotherapy; Temerty Centre for Therapeutic Brain Intervention, Centre for Addiction and Mental Health, University of Toronto, Toronto, ON, Canada; Department of Psychiatry, Temerty Faculty of Medicine, University of Toronto, Toronto, ON, Canada; Department of Child and Adolescent Psychiatry, Psychosomatics and Psychotherapy, Medical Faculty, RWTH Aachen University, Aachen, Germany

**Keywords:** schizophrenia, transcranial direct current stimulation, transcranial random noise stimulation, dorsolateral prefrontal cortex, cognition, emotion regulation

## Abstract

Cognitive deficits in schizophrenia significantly hinder functional outcomes and often remain unresponsive to conventional treatments. While initial evidence suggested potential pro-cognitive effects of electrical brain stimulation in schizophrenia, recent meta-analyses have not supported these findings, warranting further investigation on intervention optimization. This sham-controlled crossover study explored cognitive and emotional effects of bilateral dorsolateral prefrontal cortex (DLPFC) anodal transcranial direct current stimulation (tDCS) and high-frequency transcranial random noise stimulation (tRNS) in schizophrenia.

Thirty-six male patients with schizophrenia participated in a crossover trial, receiving three sessions (tDCS, tRNS, sham) in counterbalanced order with one-week intervals. tDCS and tRNS sessions involved 20-minute 2 mA anodal stimulation (tDCS) and 2 mA 100–640 Hz random noise stimulation targeting the left and right DLPFCs (F3-F4) with two extracephalic return electrodes. Executive functions (working memory, planning) were assessed during stimulation, and emotional changes were measured with the Positive and Negative Affect Schedule (PANAS) pre- and post-stimulation. Side effects and blinding efficacy were evaluated.

Both bilateral tDCS and tRNS significantly improved executive functions (i.e., problem solving) compared to sham, with tRNS additionally enhancing working memory accuracy and strategy score. Both interventions increased positive affect and reduced negative affect after the intervention, with tRNS showing greater enhancement of positive emotions. Reduced negative affect correlated with better executive functions during tRNS. Side effects were minimal, and blinding was effective for the sham condition.

Bilateral DLPFC anodal tDCS and high-frequency tRNS show promise as adjunctive treatments for schizophrenia, especially for cognitive deficits, with broader cognitive and emotional benefits observed with tRNS.

## 1. Introduction

Schizophrenia is a complex and chronic psychiatric disorder characterized by positive symptoms (e.g., hallucinations, delusions), negative symptoms (e.g., amotivation, social withdrawal), and cognitive impairments (e.g., deficits in working memory and executive functions) ^1,2^. In the latest Global Burden of Disease Study, Schizophrenia is among the top 20 diseases in 2021 in disability metrics ^3^. The early term of schizophrenia as dementia praecox ^4^ highlights the significance and relevance of cognitive deficit for the disease’s physio- and psychopathology ^5^. These cognitive deficits significantly impair daily functioning and are often resistant to conventional antipsychotic treatments, and represent one of the main obstacles to clinical and functional recovery in affected individuals ^6,7^. Up to 30% of patients with schizophrenia fail to achieve meaningful improvement with first-line antipsychotics, and even clozapine, a second-line treatment, yields response rates of only 30%-60% ^6,8^. Consequently, there is a pressing need for novel therapeutic strategies to treat schizophrenia symptoms, especially neurocognitive deficits, which are critical for improving functional outcomes in schizophrenia ^9^.

Novel treatment strategies for schizophrenia are informed by advancing knowledge of its neurobiology, including affected brain structures, functions, and networks. Functional Magnetic Resonance Imaging (fMRI) fMRI markers reveal widespread involvement in cognitive deficits, with the frontoparietal network—key for cognitive control—strongly linked to executive functions like working memory and planning, showing structural and functional abnormalities ^7^. The dorsolateral prefrontal cortex (DLPFC), a key brain region in this network, consistently shows aberrant activation during core cognitive deficits such as working memory tasks, with both hypo- and hyperactivation reported in schizophrenia ^7^. Noninvasive brain stimulation provides safe ways to modify and restore these abnormal activities in cortical and subcortical regions ^10^ and holds promise for patients with schizophrenia ^8,11^. Transcranial electrical stimulation (tES), including transcranial direct current stimulation (tDCS) and transcranial random noise stimulation (tRNS), has emerged as a promising neuromodulation tool for psychiatric disorders ^12,13^. tDCS induces sustained membrane potential shifts and ion channel modulation, with polarity-specific effects: anodal enhances excitability, cathodal reduces it ^14^. Conversely, tRNS applies random pulses across frequencies and amplitudes, leveraging stochastic resonance to amplify weak neural signals ^15^.

In schizophrenia, tES has been explored for its potential to alleviate positive, negative, and cognitive symptoms ^16,17^, with a particular focus on targeting the DLPFC due to its role in working memory, and emotion regulation ^18,19^. The DLPFC is implicated in the pathophysiology of cognitive and negative symptoms in schizophrenia, exhibiting reduced metabolism and altered connectivity ^7,20^. The rationale for using tES in schizophrenia is strengthened by the disorder’s characteristic excitation/inhibition (E/I) imbalance. Studies show that schizophrenia is associated with reduced cortical excitability, reflected in lower E/I balance as measured by EEG ^21,22^. Both tDCS and tRNS can increase E/I balance and enhance cortical excitability ^23,24^, potentially counteracting these deficits. Furthermore, tRNS combined with executive function training has increased E/I balance in ADHD ^25^, suggesting tRNS may broadly modulate neural dynamics. These findings provide a strong rationale for investigating tES as a therapeutic intervention to restore E/I balance in schizophrenia.

Early tDCS studies suggested cognitive improvements in schizophrenia via frontal connectivity changes ^26,27^, but recent meta-analyses found no robust effects on negative symptoms^28^ or cognitive outcomes ^17^, highlighting the need for optimized or novel tES protocols. In this respect, high-frequency tRNS (e.g., 100–640 Hz), has shown superior neuromodulatory effects compared to low-frequency tRNS in enhancing neuroplasticity and cognition ^29^. Despite its potential, tRNS remains underexplored in schizophrenia, with preliminary case reports suggesting benefits for negative symptoms and patient insight ^30–32^. Moreover, previous tES studies in schizophrenia have primarily utilized unilateral tDCS montages, such as anodal stimulation over the left DLPFC with a cathode over the temporoparietal cortex, showing mixed results ^17,33^. One such reason could be insufficient modulation of interhemispheric connectivity and bilateral network imbalances in the disorder ^34^. Only one tDCS study applied bilateral DLPFC anodal tDCS (with supraorbital cathodes) and reported reduced negative symptoms and cognitive gains ^35^.

The current study has two novel aspects. The use of: (1) bilateral anodal tDCS targeting both the left and right DLPFC, with return electrodes positioned on the shoulders to minimize cortical interference by the return electrodes, and (2) high-frequency tRNS using the same electrode configuration. Unlike traditional montages, the extracephalic placement of return electrodes on the shoulders aims to enhance the specificity of DLPFC stimulation by reducing unintended modulation of non-target brain regions ^36^. Furthermore, stimulating both left and right DLPFCs with the same polarity/frequency allows us to see how its modulation, regardless of laterality, which is a critical tES parameter to consider ^36^, affects cognitive and emotional performance. Additionally, we assessed emotional changes following DLPFC stimulation, given the DLPFC’s critical role in emotion regulation, which is known to be impaired in schizophrenia ^37^, and monitored emotional instability before the intervention to capture short-term mood changes that may affect cognitive outcomes ^38^.

Accordingly, the primary aim of this study was to investigate the comparable cognitive and emotional effects of bilateral DLPFC stimulation using anodal tDCS and high-frequency tRNS in patients with schizophrenia. We hypothesized that both tDCS and tRNS of bilateral DLPFCs enhance working memory and executive function, compared to sham stimulation. Furthermore, we anticipated improvements in positive affect and reductions in negative affect post-stimulation. To our knowledge, this is the first tES study that compares the effects of upregulating the bilateral DLPFC with anodal tDCS and high-frequency tRNS on cognitive deficits and emotional functioning in schizophrenia, addressing the need for optimized/novel neurostimulation interventions for the disease ^8^.

## 2. Methods

### 2.1. Study design and participants

This study used a randomized crossover design. The order of the three stimulation conditions (anodal tDCS, high-frequency tRNS, sham) was counterbalanced across participants using a Latin square design to control for order effects. Condition sequences were generated using a computer-based random sequence generator (www.random.org) by an independent researcher not involved in data collection or analysis (Figure 1). The sample size was calculated using a power analysis, and the average sample size in previous studies ^17,39^. For a repeated measures ANOVA with 3 measurements, a power of 0.90, alpha of 0.05, and a medium effect size (partial eta square of 0.06 equal to f=0.25), a minimum of 36 participants was required. Thirty-six male patients with schizophrenia (mean age = 46.11±5.36) were recruited from the inpatient schizophrenia wards in the Daroshafa Hospital for Psychiatric and Chronic Diseases (National Welfare Organization, Ardabil, Iran) (see Table for demographics). Inclusion criteria were: (1) Clinician-Rated Dimensions of Psychosis Symptom Severity ^40,41^ (Schizophrenia diagnosis was based on the DSM-5 criteria with no minimum symptom severity threshold applied), (2) being 18-50 years of age, (3) stable doses of antipsychotics [defined as a change of not more than 50% of the dose of antipsychotics) and all CNS-active medications (4–6 weeks before the experiment), and (4) the ability to provide informed written consent. Stable antipsychotic treatment was defined as no more than 50% change in dose of any medication in the 4–6 weeks prior to and throughout the study, confirmed via medical records. All participants had to meet the safety guidelines for tDCS/tRNS interventions and be fluent in their native language. Exclusion criteria included alcohol or substance dependence, history of seizures, neurological disorders, head injury, or contraindicated implants. This was a registered clinical trial (ClinicalTrials.gov Identifier: NCT06155786) approved by the Ethics Committee of Payame Noor University (Ethics code: IR.PNU.REC.1401.446). Participants and their guardians gave their written informed consent before participation. Patients were free to withdraw from the study upon request.

**Fig. 1:**
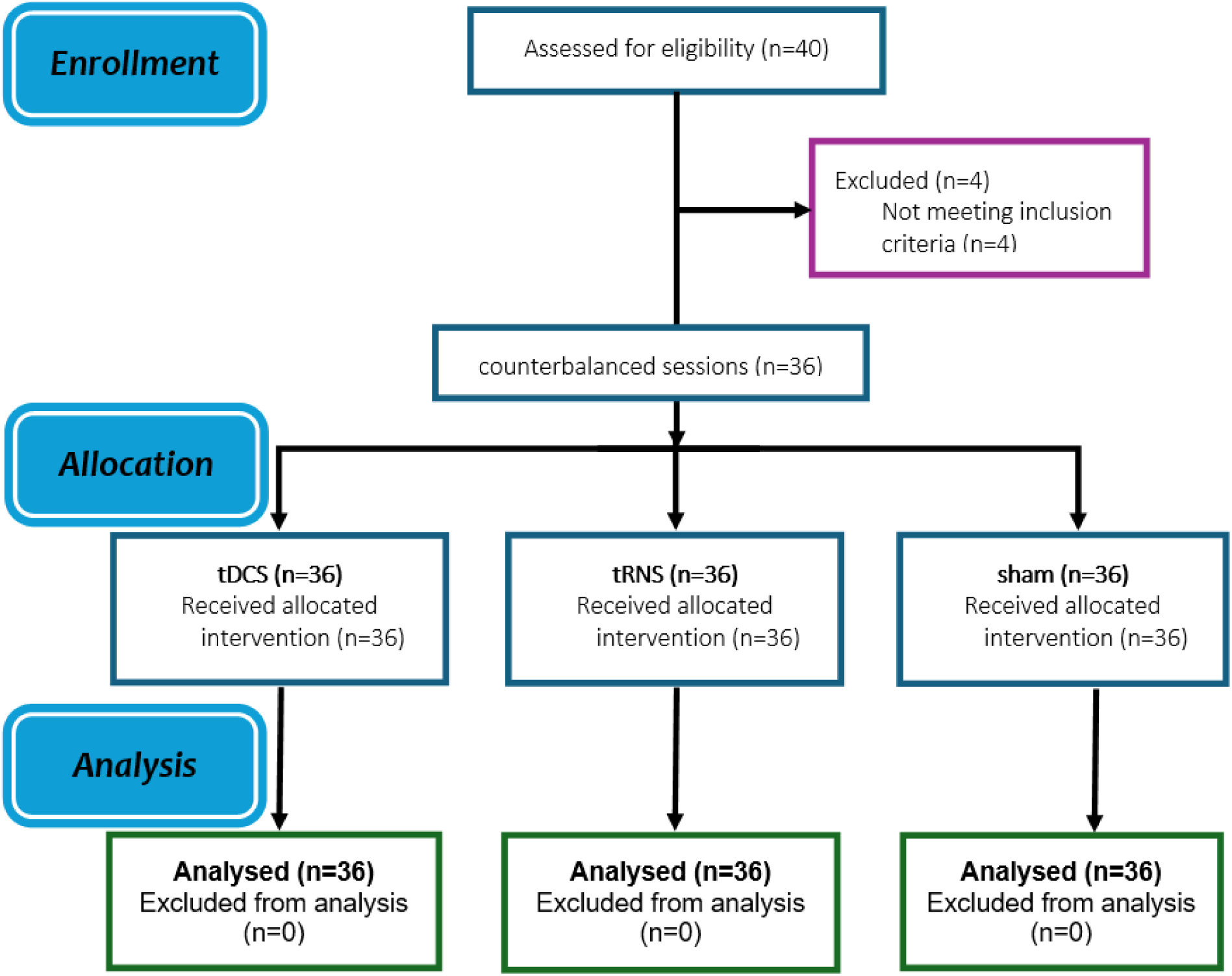
CONSORT flow diagram of study inclusion

### 2.2. Measurements

Core executive functions (working memory, planning) were assessed using two subtests of the CANTAB schizophrenia battery ^42^, including the spatial working memory task for measuring working memory and the Stockings of Cambridge task for measuring planning. Emotional changes were assessed with the Positive and Negative Affect Schedule ^43^, and tES side effects were measured with the standard survey ^44^. These measures were chosen to measure key DLPFC-dependent executive functions—working memory and planning—that are significantly impaired in schizophrenia and closely tied to functional outcomes. Although other cognitive deficits, such as attentional functioning and social cognition, are also critical within the neurocognitive domains of schizophrenia ^9,45^ they were not included in this study.

#### 2.2.1. Executive Functions Tasks

The Spatial Working Memory (SWM) and Stockings of Cambridge (SOC) tasks are designed to assess critical executive functions, specifically working memory and spatial planning. The SWM task involves a computerized setup where participants locate a yellow ‘token’ hidden in colored boxes through a process of elimination, aiming to fill an empty column. The task’s difficulty increases with up to 12 boxes, varying in color and position across trials, with outcome measures focusing on strategy scores (lower scores indicating effective strategy use) and total errors (selecting incorrect boxes). Conversely, the SOC task, derived from the Tower of London test ^46^, evaluates spatial planning and frontal lobe function by requiring participants to rearrange colored balls to match a target pattern in the fewest moves possible, progressing from simple one-move to complex multi-move problems. Outcome measures include the mean number of problems solved (higher scores better) and the mean number of moves required for problems of varying difficulty (lower scores better). Both tasks are sensitive to cognitive impairments in conditions like dementia, ADHD, and schizophrenia, providing valuable insights into executive function capabilities ^47^ (see supplementary materials for details).

#### 2.2.2. Positive and Negative Affect Schedule (PANAS)

The PANAS ^43^ was used to measure emotional states. It consists of 20 items: 10 for positive affect (e.g., “enthusiastic,” “inspired”) and 10 for negative affect (e.g., “afraid,” “irritable”). Participants rate each item on a 5-point Likert scale (1 = not at all, 5 = very much) based on their current or recent feelings. Scores for positive and negative affect are calculated separately, ranging from 10 to 50. The PANAS is recognized for its high reliability, validity, and sensitivity to short-term mood changes, making it ideal for assessing baseline emotional states and changes before and after interventions, such as tDCS, especially when targeting brain regions involved in emotional regulation (e.g., prefrontal cortex)^38^. It also provides information about whether and how subjective estimates of affect can influence performance.

### 2.3. Transcranial Electrical Stimulation

Participants underwent three sessions of tES—anodal tDCS, high-frequency tRNS, and sham stimulation—administered in counterbalanced order with a one-week interval. 2 mA stimulation was delivered using a 4-electrode device (Neurostim2, MedinaTeb Co., Iran) via 0.9% saline-soaked sponge electrodes (7×5 cm-current density of 0.06 mA/cm^2^) positioned on target regions according to the 10-20 EEG international system. In the 2 mA tDCS protocol, bilateral anodal electrodes were applied to the left DLPFC (F3) and right DLPFC (F4), with cathodes over the right and left shoulders. The tRNS protocol used the same electrode placement with target electrodes over the left and right DLPFCs (F3-F4) and two return electrodes over the left and right shoulders, an intensity of 2 mA base to peak (without DC offset), a frequency of 100–640 Hz, a stimulation was applied for 20 minutes. The frequency range was selected based on the results of previous studies that showed neuromodulatory effects of 100–640 Hz tRNS compared to 0–100 Hz tRNS ^48^ although induced electrical fields are independent of stimulation frequency in tRNS ^29^. For sham stimulation, the same electrode configuration was used, featuring a 30-real stimulation with a 30-second ramp-up at the start and a 30-second ramp-down at the end. Half of the sham sessions involved the application of direct current, while the other half used random noise to simulate both stimulation types, evenly distributed across the patients. For the remaining 18.5 minutes, the device was turned off without the knowledge of the patients. In all sessions, both electrodes were positioned longitudinally along antero-posterior axis of the target regions to ensure a 6 cm distance between electrodes and were fixed with headbands. Five minutes after starting the stimulation, participants performed the SWM and SOC tasks. After each stimulation session, participants completed a side-effect checklist ^44^ and were asked to guess about the stimulation type (active or sham) for assessing blinding efficacy. The Report Approval for Transcranial Electrical Stimulation (RATES) checklist ^49^ of the intervention and study design is presented in Table 2.

**Table 1.** Baseline sociodemographic and clinical characteristics of the participants

| Characteristics | (N = 36) |
| --- | --- |
| Schizophrenia/schizoaffective disorder | 23/13 |
| Age, years | 46.11 ± 5.36 |
| Education level, years | 12.46 ± 3.68 |
| BMI, kg/m<sup>2</sup> | 28.63 ± 2.74 |
| Weekly regular exercise, hours | 1.26 ± 0.53 |
| Handedness (right-left) | 32/4 |
| Smoker, n (%) | 12 (33.33) |
| Hypertension, n (%) | 5 (13.88) |
| Diabetes mellitus, n (%) | 2 (5.55) |
| Onset age, years | 21.68 ± 6.22 |
| Length of illness, years | 16.78 ± 7.25 |
| Antipsychotic dosage, mg/day<sup>a</sup> | 17.38 ± 8.27 |
| Anticholinergic dosage, mg/day<sup>b</sup> | 0.86 ± 1.89 |
| Sedative-hypnotic dosage, mg/day<sup>c</sup> | 38.75 ± 38.43 |
| PANSS total score | 56.58 ± 14.56 |
| PANSS negative symptoms subscale score | 16.06 ± 6.23 |

**Table 2:** RATES (Report Approval for Transcranial Electrical Stimulation) checklist ^49^

|  |  |  |  |  |
| --- | --- | --- | --- | --- |
| Participants | Sample Size (Dx), Recruitment Process, Medication | | 36 male Inpatients with schizophrenia, recruited from Daroshafa Hospital (Ardabil, Iran), stable doses of antipsychotics (change $\leq 50\%$ in 4–6 weeks before experiment) | |
| | Age(yrs), Sex(F:M), Edu(yrs), Handedness(R:L:D) | | 46.11 ± 5.36, 0:36, n/s, adjustable for left and right-handed. see Table 1 | |
|  | Hours of Sleep, Consumption of Caffeine, Nicotine, and Alcohol |  | Under no sleep pressure (2:00–5:00 p.m.), n/s, allowed, n/s |  |
|  | Participant Eligibility Criteria: Aged 18–50 years, schizophrenia diagnosis per Clinician-Rated Dimensions of Psychosis Symptom Severity, stable antipsychotics, ability to provide informed consent. Exclusions: alcohol/substance dependence, seizures, neurological disorders, head injury, contraindicated implants. Fluent in native language, normal or corrected-to-normal vision. |  |  |  |
| Stimulator | Stimulator: Neurostim2 (MedinaTebCo., Iran), 4-electrode device for tDCS and tRNS |  |  |  |
|  | Sham Option | Yes | Waveform | Constant direct current (tDCS); random noise 100–640 Hz (tRNS) |
|  | Output Channels | 4 channels | Stimulator Safety Features | The devices had current and voltage limits |
|  | Current Resolution | n/s | Monitoring and Feedback | n/s |
| Electrodes | Positioning | International 10-20 EEG system | Inter-electrode Distance | At least 6 cm |
|  | Shape | Rectangular | Assembly | Fixed with headbands |
| | Size | 35 cm<sup>2</sup> (7×5 cm) | Contact Medium | 0.9% saline-soaked sponge |
|  | Orientation | longitudinal along the medio-lateral axis (conventional) | Impedance | n/s |
|  | Material | Sponge electrodes | Connector Position | n/s |
|  | Number: Four electrodes (two anodal/target over F3–F4, two return on shoulders) |  |  |  |
|  | Montage: Bilateral DLPFC: targets at left (F3) and right (F4) DLPFC, return electrodes on left and right shoulders (extracephalic). Same for tDCS, tRNS, sham. |  |  |  |
| Current | Intensity (mA) | 2 mA | Amplitude | Peak to zero (tDCS); 2 mA base to peak, no DC offset (tRNS) |
|  | Density (V/m) | 0.06 mA/cm<sup>2</sup> | Personalization | None |
|  | Distribution (Method) | n/s | Duration (min) | 20 min |
|  | Frequency (Hz) | n/a (tDCS); 100–640 Hz (tRNS) | Ramp up/down (sec) | 30:30 |
|  | Polarity | Anodal (left DLPFC excitatory); cathodal (right DLPFC inhibitory) | Warm-up time (min) | n/a |
|  | Waveform | Constant DC (tDCS); random noise (tRNS) | Sham Characteristics | 30 sec real stimulation (half DC, half noise), then off for 18.5 min, with 30 sec ramp up/down |
| Procedure | Study Setting and Site | Dedicated research space at the hospital | Attrition (n) | 0 |
|  | Hypothesis Statement | Exploratory | Blinding Method | Double-blind |
|  | Preregistration | Registered (ClinicalTrials.gov NCT06155786) | Ethical Considerations | Approved by the Ethics Committee (IR.PNU.REC.1401.446) |
|  | Session Duration (min) | 20 min stimulation + tasks (~15 min during stim) | Safety Monitoring | Side effects checklist post-session |
|  | Total Number of Sessions | 3 sessions (tDCS, tRNS, sham) | Informed Consent Process | yes |
|  | Session Frequency | One-week intervals | Conflict of Interest | None reported |
|  | Concurrent Intervention | No (stable meds only) | tES operator | Trained tES operator |
|  | Randomization (Method) | Counterbalanced order | Data Analysis Plan | Repeated measures ANOVA, correlations |
|  | Counterbalancing | Yes (counterbalanced order) | Data Availability | yes |
|  | Study Design: Randomized, sham-controlled, crossover |  |  |  |
|  | Stimulation and Assessment Task Timing: Online (tasks started 5 min into stimulation) |  |  |  |
|  | Inter-session Interval: One week |  |  |  |
|  | Data Collection Time Points: Pre- and post-stimulation (PANAS), during stimulation (cognitive tasks), post (side effects, blinding) |  |  |  |
|  | Baseline Assessment: Yes (pre-PANAS, eligibility) |  |  |  |
|  | Control Intervention: Sham (30 sec ramp up, 30 sec stim, 30 sec ramp down, then off) |  |  |  |
|  | Outcome Measure: Cognitive (SWM, SOC from CANTAB), emotional (PANAS), side effects, blinding efficacy |  |  |  |

### 2.4. Procedure

Before the experiment, participants completed a brief questionnaire to evaluate their suitability for brain stimulation. All participants received 3 sessions of tES with one-week intervals between the sessions. To avoid confounding effects of the intervention at circadian non-preferred time and sleep pressure which can significantly affect neuroplasticity induction and cognitive performance ^50,51^, all stimulation sessions took place at a specific time of day between 2:00 and 5:00 p.m. Cognitive measures were evaluated *during* each 20-min stimulation session (online), started 5 minutes after the beginning of the stimulation and took about 15 minutes to complete. This timing allowed the tasks to be performed while cortical excitability was actively modulated, enabling the assessment of state-dependent effects of tES on cognitive performance, rather than post-stimulation aftereffects, and is in line with previous studies ^52–54^. To assess changes in affective states during the experimental procedure, participants completed the PANAS twice throughout each session, before and after each stimulation. This was also done to control for baseline emotional variability ^38^.

All patients had normal or corrected-to-normal eyesight. Patients were instructed about the tasks before the beginning of the experiment. Stimulus presentation in all computerized tasks was conducted by a 15.6” screen, at a viewing distance of approximately 50 cm, and the response box was adjustable for left and right-handed individuals. To mitigate potential practice effects and stabilize performance, stimuli were presented in a randomized order for each run, and a practice session familiarized participants with the task before the baseline. Practice session data were excluded from the analysis. None of the patients received any type of psychotherapy during the study. All sessions were conducted in a dedicated space for research with similar environmental conditions in all sessions. Participants were blind to the study hypotheses and stimulation conditions. To maintain a double-blind design, a separate investigator prepared the device and administered the stimulation while the experimenter who evaluated emotional changes (PANAS) and conducted the outcome measures during stimulation was blinded to the tES conditions (Figure 2).

**Figure 2:**
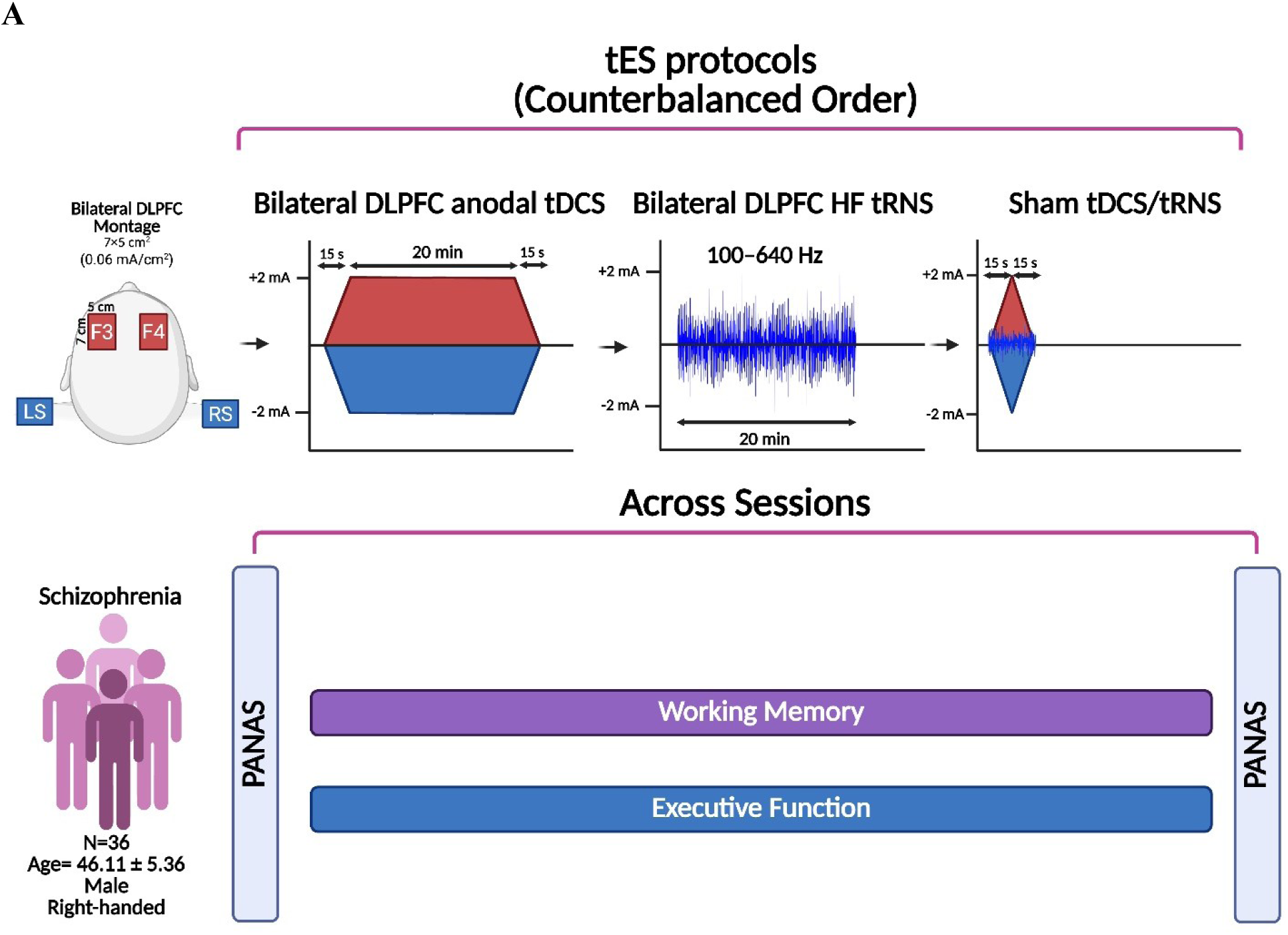
Experimental procedure. This study was a randomized, double-blind, crossover. Participants were randomly assigned to three tES sessions with at least a one-week interval: bilateral DLPFC anodal tDCS, bilateral DLPFC high-frequency tRNS, and sham tES. In all tES sessions, 4 electrodes were used with active electrodes applied to the left DLPFC (F3) and right DLPFC (F4), with return electrodes over the right and left shoulders. Participants underwent pre-and post-intervention assessments of emotional states. Additionally, executive functions (working memory and planning) were measured with the Cambridge Neuropsychological Test Automated Battery (CANTAB) during each stimulation session. After each stimulation session, participants completed a side-effect checklist and were asked to guess about the stimulation type for assessing blinding efficacy. Cognitive measures were evaluated during each 20-minute stimulation session (online), started 5 minutes after the beginning of the stimulation, and took about 15 minutes to complete.

### 2.5. Statistical Analysis

Data analyses were conducted with the statistical package SPSS, version 26.0 (IBM, SPSS, Inc., Chicago, IL), and GraphPad Prism 8.2.1 (GraphPad Software, San Diego, California). The normality and homogeneity of data distribution and variance were confirmed by Shapiro-Wilk and Levene tests, respectively. To explore the effect of tES conditions on cognitive performance (working memory, planning), a repeated measure ANOVA was conducted with “stimulation” (Sham, tDCS, tRNS) as the within-subject factor. Mauchly’s Test of Sphericity was employed to assess the sphericity of the data, and adjustments to degrees of freedom were made using the Greenhouse-Geisser method when necessary. In the event of significant findings in the ANOVAs, post-hoc analyses were executed using Bonferroni-corrected t-tests to account for multiple comparisons. The scores on PANAS before and after each stimulation condition were analyzed with a 3 (stimulation) × 2 (emotion) × 2 (time) factorial ANOVA, and baseline PANAS scores were analyzed with a one-way ANOVA. Two-sided Pearson correlations examined relationships between tES effects on task performance and mood changes (PANAS) only when Bonferroni-corrected pre-post PANAS differences were significant, limiting tests to hypothesis-relevant cases and minimizing Type I error. These exploratory correlations between cognitive performance and mood changes were not further corrected because they were conditional on significant pre-post PANAS differences and were few in number. Side effects were analyzed with the same one-way ANOVAs for each domain. Blinding efficacy was examined with the chi-squared test for Independence on participants’ guesses of blinding (0, 1) and Bang’s Blinding Index (–1, 1) ^55^, with negative values suggesting that participants frequently guessed the opposite of their actual treatment. The critical level of significance was 0.05 for all statistical analyses.

## 3. Result

### 3.1. Safety outcomes and blinding

The stimulation was well-tolerated without major side effects in all participants, and none dropped out of the study. Minor side effects were mild (mean ratings < 1.0 on a 0–4 scale) and did not differ significantly between conditions (Table 3). One-way ANOVAs revealed non-significant effect of stimulation on pain (*F*_2,70_=0.761, *p*=0.470), itching (*F*_2,70_=2.369, *p*=0.099), fatigue (*F*_2,70_=0.583, *p*=0.560), burning sensation (*F*_2,70_=0.957, *p*=0.387), skin redness (*F*_2,70_=1.467, *p*=0.235), and problems in concentration (*F*_2,70_=0.062, *p*=0.940), indicating that side effects did not differ between tES conditions (Table 3). Regarding blinding efficacy, four patients during sham tES and one patient during tDCS and tRNS guessed no stimulation (chose 0). The results of the Chi-Square test (χ2=3.176; p=0.204) and Bang’s Blinding Index (tDCS and tRNS, BBI = 0.94 each, sham BBI = -0.77) indicated that blinding for the sham condition was successful (negative BBI reflects effective sham credibility) ^56^, suggesting that participants’ awareness of their tDCS condition did not significantly influence their perception of side effects.

**Table 3.** Mean and standard deviation of reported side effects and outcome measures, along with ANOVA results for the side effects

| Measures | Stimulation condition, M (SD) |  |  | Statistics |  |  |
| --- | --- | --- | --- | --- | --- | --- |
|  | Sham | tDCS | tRNS | *df* | *F* | *p* |
| **Pain** | 1.06 (0.23) | 1.08 (0.28) | 1.14 (0.35) | 2,70 | 0.761 | 0.470 |
| **Itching** | 1.11 (0.32) | 1.31 (0.52) | 1.36 (0.11) | 2,70 | 2.369 | 0.099 |
| **Fatigue** | 1.25 (0.44) | 1.31 (0.48) | 1.19 (0.40) | 2,70 | 0.583 | 0.560 |
| **Burning** | 1.11 (0.46) | 1.25 (0.44) | 1.12 (0.38) | 2,70 | 0.957 | 0.387 |
| **Skin Redness** | 1 (0.00) | 1.08 (0.28) | 1.06 (0.23) | 2,70 | 1.467 | 0.235 |
| **Focus problem** | 1.17 (0.38) | 1.17 (0.38) | 1.19 (0.40) | 2,70 | 0.062 | 0.940 |
| **Cognitive outcome measures** |  |  |  |  |  |  |
| **SWM error** | 42.78 (13.05) | 36.92 (10.55) | 34.83 (13.61) | - | - | - |
| **SWM strategy** | 38.06 (3.8) | 39.69 (3.77) | 37.75 (4.03) | - | - | - |
| **SOC- mean problem solved** | 4.61 (2.06) | 7.42 (1.25) | 7.53 (1.95) | - | - | - |
| **SOC- 2 moves** | 2.78 (1.31) | 2.47 (1.03) | 2.76 (1.02) | - | - | - |
| **SOC - 3 moves** | 4.6 (1.06) | 3.18 (0.5) | 3.28 (0.85) | - | - | - |
| **SOC - 4 moves** | 6.68 (0.93) | 5.02 (0.85) | 5.97 (1.51) | - | - | - |
| **SOC - 5 moves** | 6.61 (2.21) | 5.86 (1.11) | 6.53 (1.52) | - | - | - |

tDCS = transcranial Direct Current Stimulation, tRNS = transcranial Random Noise Stimulation, M = mean, SD = Standard Deviation, SWM = Spatial Working Memory, SOC = Stockings of Cambridge,

### 3.2. tES effects on executive functions

A repeated-measure ANOVA was conducted to investigate the effect of stimulation on cognitive performance across outcome measures. For working memory, a significant main effect of stimulation (F_2,70_=3.479, p=.036, *ηp^2^*=0.090) was observed on SWM strategy score (lower strategy score = efficient performance). Bonferroni-corrected post-hoc t-tests revealed a lower strategy score during tRNS when compared with tDCS (MD=1.944, *p*=0.022) but not with the sham condition, despite a numerically larger score, *Figure B*. For SWM error, the analysis revealed a significant main effect of stimulation (*F*_2,70_=4.060, *p*=0.021, ηp^2^=0.104). Bonferroni-corrected post-hoc t-tests yielded significantly fewer errors only during tRNS compared with sham (MD=7.944, *p*=0.028) and not during tDCS, *Figure A*.

For the planning measured by SOC, the ANOVA results for the mean number of problems solved indicated a significant main effect of stimulation (*F*_2,70_=34.309, *p*≤0.001, ηp^2^=0.495). Bonferroni-corrected post-hoc t-tests yielded a higher number of solved problems on minimum move solutions during both tDCS (MD=2.806, *p*≤0.001) and tRNS (MD=2.917, *p*≤0.001) stimulations compared to the sham condition, *Figure A.* Results for 2-moves (*F*_2,70_=9.15, *p*=0.405, ηp^2^= 0.025) and 5-moves difficulty (F_2,70_=2.346, p=0.103, ηp^2^= 0.063) showed a non-significant main effect of stimulation. In the 3-moves (F_2,70_=29.919, *p*≤.001, ηp^2^= 0.461) and 4-moves (F_1.55,54.18_=19.641, *p*≤0.001, ηp^2^= 0.359) difficulty conditions, however, a significant main effect of stimulation was found indicating that patients under bilateral DLPFC tDCS (3 and 4 moves) and tRNS (3-moves) needed lower number of moves to complete the task (i.e., more efficient problem solving). Bonferroni-corrected post-hoc t-tests revealed better performance during tDCS (MD_3-moves_=1.417, *p*≤0.001; MD_4-moves_=1.661, *p≤*0.001) and tRNS (MD_3-moves_=1.319, *p*≤.001; MD_4-moves_=.951, *p*=0.005) compared with sham *Figure D-G*.

### 3.3. Emotional stability and tES effects on positive/negative affect

PANAS was used to (1) assess patients’ emotional stability before each stimulation (comparing only pre-intervention scores) and (2) monitor emotional changes post-stimulation (comparing pre- and post-intervention scores). There were no significant differences in pre-intervention PANAS positive (*F*_2,70_=0.812, *p*=0.448) and negative scores (*F*_2,70_=1.483, *p*=0.234) across conditions, indicating a similar emotional state among patients before each stimulation. With respect to stimulation-specific effects, the results of the 3×2×2 ANOVA showed a significant three-way interaction of stimulation×time×emotion on PANAS scores (*F*_2,70_=23.053, *p*<0.001, *ηp^2^*=0.397) as well as stimulation×time (*F*_1,35_=8.153, *p≤*0.001, *ηp^2^*=0.189) and time×emotion (*F*_1,35_=34.492, *p*<0.001, *ηp^2^*=0.496) interactions. The main effect of time (*F*_1,35_=119.73, *p*<0.001, *ηp2=0.774*) was significant too. For within-condition comparisons, the Bonferroni-corrected post hoc t-tests showed that positive emotions significantly increased after both the tDCS (*p*<0.001) and tRNS (*p*<0.001) conditions, but not after sham tES (*p*=0.190), while negative emotions significantly decreased after tDCS (*p*<0.001) and tRNS (*p*<0.001), but not after sham tES (*p*=0.146). Post hoc analyses of the between-condition comparisons revealed that both tDCS (*p*<0.001) and tRNS (*p*<0.001) significantly reduced negative emotions after the intervention compared to the sham condition, whereas only tRNS (*p*=0.003) but not tDCS (*p*=0.108) significantly enhanced positive emotions post-intervention relative to the sham (Figure 4AB).

**Figure 3.**
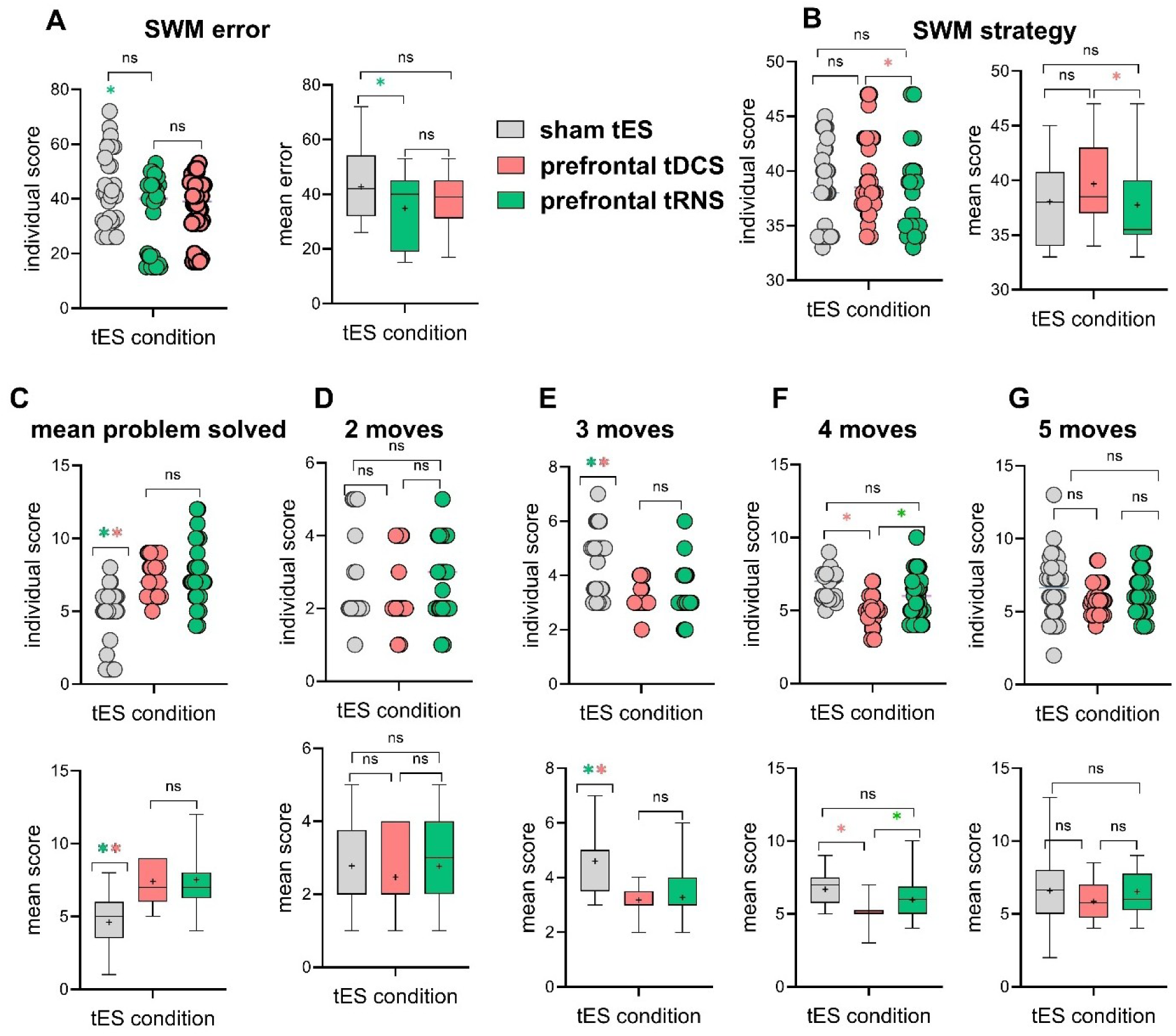
Effect of tES on working memory (**A, B**) and executive functions (**C-G**) in 36 patients with schizophrenia. *Note*: SWM: spatial working memory, SOC: Stockings of Cambridge. * denotes significant differences between active tES conditions (red for tDCS, green for tRNS) versus sham or other conditions, if applicable, at p < 0.05 (Bonferroni corrected) based on pairwise comparison results. Brackets indicate comparisons between active conditions. Error bars, where applicable, represent standard error of the mean (s.e.m.). **ns**: non-significant.

**Figure 4:**
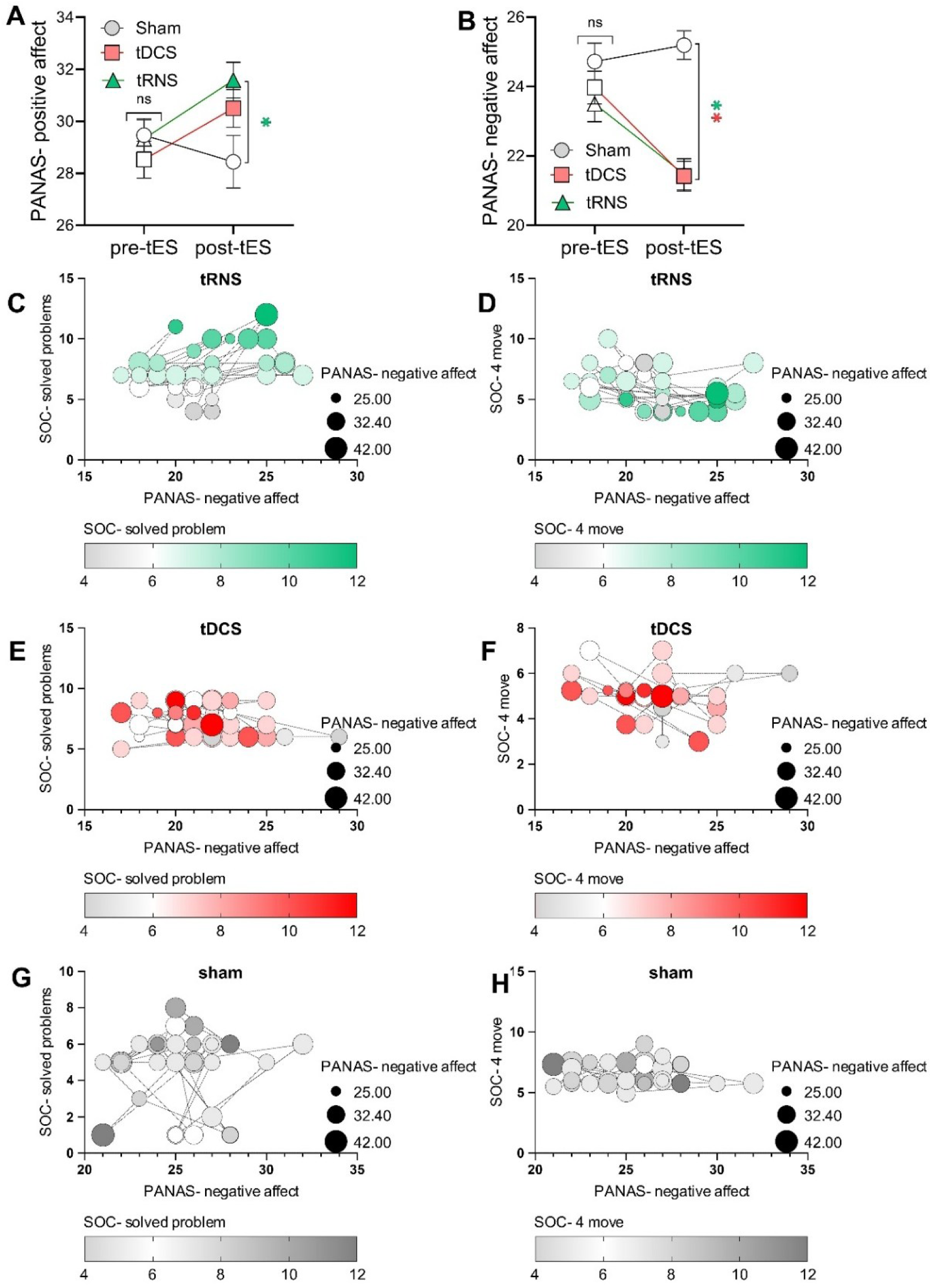
The effects of tES sessions on positive (A) and negative (B) emotional affect, along with their association with cognitive function (C-H). Both anodal bilateral DLPFC tDCS and bilateral DLPFC tRNS, but not sham tES, significantly increased positive affect and decreased negative affect post-intervention. Note: only significant correlations are presented. Figures **A-B** show that emotional stability before the intervention showed no significant differences across stimulation conditions, while both tDCS and tRNS significantly reduced negative affect and also increased positive affect. Filled symbols indicate a significant difference between post- and pre-intervention, while [*] denotes a significant difference compared to sham. Figures C-H present scatterplots of patients’ post-intervention cognitive performance and negative affect, showcasing only significant comparisons. In C, an increase in solved problems after the intervention (represented by darker green nodes) correlates with higher PANAS-negative affect scores (closer to 15), suggesting that improved executive function is associated with reduced negative affect. In D, a lower number of moves required to solve problems (darker green nodes) correlates with lower PANAS-negative affect scores (closer to 15), indicating that better performance in the SOC task may also be linked to reduced negative affect. Scatterplots E-F and G-H depict similar data but show no significant results for the tDCS and sham conditions.

### 3.4. Correlational analysis

The results of Pearson’s correlational analyses of the association between cognitive performance and mood changes revealed a significant positive correlation between reduced negative affect (i.e., a greater decrease in negative emotions post-intervention, indicating improved emotional state) and the mean value of solved problems during tRNS (r=0.387, *p_two-sided_*=0.020) (Fig. 4C) indicating that higher solved problems was associated with less negative affects. Additionally, a significant negative correlation was found between the mean moves required to solve the problem at level 4 difficulty (lower values better) and decreased negative affect during tRNS (r= -0.363, *p_two-sided_*=0.029) (Fig. 4D), indicating that the fewer number of moves for problem solving was associated with lower negative affect. The remaining correlations (Fig. 4E-H) were non-significant.

## 4. Discussion

This randomized sham-controlled study explored the cognitive and emotional effects of bilateral DLPFC anodal tDCS and high-frequency tRNS in patients with schizophrenia. Our results show that both active tES interventions enhanced executive function measured via a problem-solving task, with tRNS additionally improving working memory performance accuracy. Additionally, both tDCS and tRNS increased positive affect and decreased negative affect after the intervention, with tRNS showing a more pronounced effect on positive emotions. These findings underscore precognitive effects and have implications with respect to considerations regarding the therapeutic potential of bilateral DLPFC excitatory stimulation as an adjunctive treatment for schizophrenia.

### 4.1. Cognitive Effects of Bilateral DLPFC tDCS and tRNS

In this study, both DLPFC tDCS and tRNS improved executive function, as shown by increased problem-solving scores and fewer moves required in the executive functions task compared to sham stimulation. Furthermore, tRNS demonstrated additional benefits in working memory by improving both strategy (numerically) and errors (significantly vs. sham), while tDCS showed less efficient strategy vs. tRNS only, though the difference was not significant with the sham. This cognitive improvement aligns with the DLPFC’s critical role in executive processes and its known dysfunction in schizophrenia ^7,9,57^ and indicates that upregulating bilateral DLPFC activity enhances core cognitive deficits, consistent with prior evidence of prefrontal hypoactivity contributing to cognitive deficits in schizophrenia ^7,20,58,59^. Additionally, this supports the potential cognitive benefits of tES in schizophrenia, specifically the tRNS, which were not demonstrated in a recent meta-analysis of tDCS trials ^17^. Although these cognitive effects were robust, the large effect sizes (ηp² > .45) should be interpreted with caution due to the potential for residual practice effects in this crossover design, despite randomization and washout procedures. Nevertheless, this should not diminish the validity of the intervention’s impact on the observed results.

A key distinction in our study is stimulation of bilateral DLPFC, a method applied in only one tDCS study in Sawfi’s meta-analysis ^17^, which also demonstrated improvements in negative symptoms and cognitive deficits ^35^. These findings further support the notion that upregulation of both left and right DLPFCs is more effective than unilateral upregulation, as observed in the majority of tES studies on schizophrenia. This suggests that bilateral DLPFC excitatory tES techniques (i.e., anodal tDCS, high-frequency tRNS) may provide a more effective approach to addressing core cognitive deficits in schizophrenia—a critical therapeutic target that is not targeted by standard treatments ^6,7^. Furthermore, this protocol configuration (with extracranial return electrodes) emphasizes the importance of target specificity in tES studies, which is often compromised by placing the cathode over regions that are essential for both cognitive and emotional functioning (e.g., supraorbital, frontopolar cortex, contralateral PFC), yet their role is ignored.

### 4.2. Emotional Effects and Relevance to DLPFC Upregulation

We also found that both bilateral DLPFC anodal tDCS and high-frequency tRNS reduced negative affect after the stimulation compared to the sham, with tRNS also enhancing positive emotions compared to the sham. These effects are consistent with the DLPFC’s role in emotion regulation, where it exerts top-down control over limbic regions to modulate affective responses ^18,60^. They are further in line with previous tES studies that have targeted DLPFC and shown mood/emotional enhancement in schizophrenia, including negative symptoms ^61–63^, and other emotional disorders with DLPFC abnormalities ^64–68^. The more pronounced emotional effect of tRNS may reflect its greater capacity to enhance prefrontal excitability, potentially amplifying its regulatory influence on emotional processing. Furthermore, our correlational analysis revealed that reduced negative affect during tRNS was associated with better executive function, suggesting an interplay between emotional and cognitive improvements. This finding supports a bidirectional relationship between cognition and emotion in schizophrenia, where reduced negative affect may facilitate executive function by alleviating emotional interference on prefrontal resources, or enhanced cognition may improve emotion regulation via modulated DLPFC activity ^18,38^.

Another point to consider here is that while both interventions reduced negative affect compared to sham, only tRNS significantly enhanced positive affect, suggesting tRNS may exert broader emotional benefits. This could be associated with tRNS’s unique stochastic resonance mechanism, which amplifies weak neural signals and enhances prefrontal synchronization more robustly than tDCS’s polarity-dependent excitability shifts ^15,29^. Additionally, bilateral prefrontal Trns is shown to increase E/I marker in the brain ^24^. By improving E/I balance and neural dynamics disrupted in schizophrenia ^21,22,34^, tRNS might facilitate greater top-down regulation of positive affective states via DLPFC-limbic pathways ^18^. These findings underscore tRNS’s potential as a more versatile tool for addressing the full spectrum of emotional dysregulation in schizophrenia, warranting further mechanistic studies with neuroimaging to delineate these effects.

### 4.3. Mechanisms of tDCS and tRNS

The distinct mechanisms of tDCS and tRNS, although not yet fully understood, might explain the partially differing effects between active interventions. tDCS delivers a constant current that shifts neuronal resting membrane potentials, with anodal stimulation increasing excitability ^10^. In schizophrenia, anodal tDCS over the DLPFC is thought to counteract hypofrontality, enhancing prefrontal activity to support cognitive and emotional functions ^16,28^. However, anodal tDCS effects are related to excitability enhancement and do not affect brain oscillatory activities, and may be less effective for tasks requiring rapid neural adaptability, such as working memory ^69^. Conversely, tRNS applies random oscillatory currents within a high-frequency range (100–640 Hz), enhancing both excitability and synchronization, promoting stochastic resonance—a process where noise enhances weak signals in neural systems ^29,70^. This mechanism may improve signal-to-noise ratios and neuronal synchronization, which are disrupted in schizophrenia ^34,71^. This mechanism may further modulate E/I balance by improving signal-to-noise ratios and neuronal synchronization, which are disrupted in schizophrenia ^21,22^. EEG studies suggest that tRNS increases E/I balance, potentially more effectively than tDCS, by enhancing cortical excitability and facilitating neuroplasticity, which aligns with its observed benefits in conditions like ADHD when combined with cognitive training ^25^. High-frequency tRNS has been shown to increase cortical excitability and facilitate neuroplasticity more effectively than tDCS ^48^, potentially explaining its broader cognitive benefits, particularly in working memory. These mechanistic differences, particularly the modulation of E/I balance as a core mechanism for both tDCS and tRNS, highlight tRNS as a promising tool for modulating the complex neural dynamics of schizophrenia.

### 4.4. Implications for Novel tES Treatment in Schizophrenia

Our findings have implications for advancing excitatory tES as a novel treatment for schizophrenia. The use of bilateral DLPFC stimulation (via anodal tDCS or high-frequency tRNS) with extracephalic return electrodes could be introduced as an innovative tES montage that enhances target region specificity by minimizing unintended effects on non-target regions ^36^ although it should be replicated in trials with a larger sample size. This approach may optimize modulation of the frontoparietal network, crucial for cognitive control ^7,69^. Although preliminary, these findings warrant confirmation in larger, multi-site studies to establish their potential contribution toward standardizing tES protocols in schizophrenia. The pro-cognitive and emotional effects of bilateral DLPFC tRNS and tDCS, specifically tRNS, may suggest their application as an adjunctive therapy, particularly for cognitive deficits in schizophrenia. Its emotional benefits, if proven in other trials, further broaden its therapeutic scope, addressing affective symptoms often neglected in current treatments ^37^.

### 4.5. Limitations, strengths, and future Directions

This study’s limitations include the absence of neuroimaging or neurophysiological data, which could have offered more detailed insights into the intervention’s neurophysiological effects. Additional cognitive assessments, which are central to neurocognitive domains in schizophrenia (e.g., attentional functioning and social cognition ^9,45^), are also recommended for future studies. These assessments are recommended to be conducted after the intervention to capture the effects of tES. Second, the study included only male participants, which may limit the generalizability of the results ^72^. Future studies should therefore include both sexes to determine whether stimulation effects differ by sex and also consider measuring tDCS aftereffects in addition to online effects. While randomized stimuli, performance stabilization training, and inter-session intervals minimized practice effects, these remain potential confounds. Future studies should also consider unified sham protocols, as splitting DC and random noise could subtly influence blinding. Finally, emotional changes were solely assessed via self-report PANAS. Future research should include physiological or implicit measures to validate affective modulation. Despite these limitations, the study’s strengths include its innovative use of bilateral DLPFC stimulation with extracephalic return electrodes, enhancing targeting specificity, and its sham-controlled, crossover design, which minimized individual differences and ensured robust comparisons between tDCS, tRNS, and sham conditions and finally, the inclusion of blinding efficacy, which is missed in the majority of tES studies in schizophrenia. Future studies should investigate the durability of these effects with repeated sessions and optimize stimulation parameters (e.g., intensity, duration) to maximize outcomes ^73^. Additionally, integrating neuroimaging or electrophysiological measures could clarify the neural mechanisms of tES, paving the way for personalized treatment strategies in schizophrenia.

## Data Availability Statement

The data that supports the findings will be publicly available at OSF and will be accessible after the publication of the study.

## Acknowledgments

none.

## Credit Author Contributions

**EJ**: Investigation, Project administration, Data curation, Validation. **AM:** Resources. **ZV**: Formal analysis (lead), Writing - Original draft (support), Visualization. **AA**: Software, Investigation (Cognitive assessment), Project administration (patient diagnosis and recruitment). **AFJ**: Visualization, Modeling. **RCK:** Writing - review & editing. **MAN:** Writing - review & editing. **DMB:** Writing - review & editing. **MAS**: Conceptualization, Methodology, Supervision, Formal analysis (support), Writing - Original draft, Writing - Review and Editing.

